# COVID-19 vaccination acceptability in the UK at the start of the vaccination programme: a nationally representative cross-sectional survey (CoVAccS – wave 2)

**DOI:** 10.1101/2021.04.06.21254973

**Authors:** Susan M. Sherman, Julius Sim, Megan Cutts, Hannah Dasch, Richard Amlôt, G James Rubin, Nick Sevdalis, Louise E. Smith

## Abstract

**Aim:** To investigate factors associated with intention to have the COVID-19 vaccination following initiation of the UK national vaccination programme.

**Methods:** 1,500 adults completed an online cross-sectional survey (13^th^–15^th^ January 2021). Linear regression analyses were used to investigate associations between intention to be vaccinated for COVID-19 and sociodemographic factors, previous influenza vaccination, attitudes and beliefs about COVID-19, attitudes and beliefs about COVID-19 vaccination and vaccination in general. Participants’ main reasons for likely vaccination uptake/decline were also solicited.

**Results:** 73.5% of participants (95% CI 71.2%, 75.7%) reported being likely to be vaccinated against COVID-19, 17.3% were unsure (95% CI 15.4%, 19.3%), and 9.3% (95% CI 7.9%, 10.8%) reported being unlikely to be vaccinated. The full regression model explained 69.8% of the variance in intention. Intention was associated with having been/intending to be vaccinated for influenza last winter/this winter, and with stronger beliefs about social acceptability of a COVID-19 vaccine; the need for vaccination; adequacy of information about the vaccine; and weaker beliefs that the vaccine is unsafe. Beliefs that only those at serious risk of illness should be vaccinated and that the vaccines are just a means for manufacturers to make money were negatively associated with vaccination intention.

**Conclusions:** Most participants reported being likely to get the COVID-19 vaccination. COVID-19 vaccination attitudes and beliefs are a crucial factor underpinning vaccine intention. Continued engagement with the public with a focus on the importance and safety of vaccination is recommended.

## INTRODUCTION

One year on from the emergence of COVID-19 in China in December 2019, there have been more than 112 million cases of COVID-19 and nearly 2.5 million deaths worldwide [1]. While countries have implemented a variety of public health measures to try to prevent the spread of the virus, scientists across the world have worked on developing effective vaccines. On 2nd December 2020, the United Kingdom (UK) became the first country to approve a COVID-19 vaccine that had been through a large-scale trial [2] and on the 8th December, the first dose of the Pfizer/BioNTech vaccine was administered [3]. This was swiftly followed by UK approval of the Oxford/AstraZeneca vaccine on 30th December 2020 and the Moderna vaccine on 8th January 2021. Given the severity of the pandemic and associated clinical outcomes, it is imperative that COVID-19 vaccination uptake is maximized so that, alongside ongoing protective public health practices, the spread of infection can be reduced [4]. To achieve this, we need to understand the factors that affect people’s willingness to have a vaccine.

The existing peer-reviewed research exploring the acceptability of a COVID-19 vaccination was all conducted before a vaccination was available [e.g. 5, 6, 7 from the UK, 8 for a systematic review of global acceptance rates], when details about the actual vaccination were still a matter of speculation. For example, in a survey of 1500 UK adults that we conducted in July 2020 [5], 64% of participants reported being very likely to be vaccinated against COVID-19, 27% were unsure, and 9% reported being very unlikely to be vaccinated. Intention to be vaccinated was associated with: more positive general COVID-19 vaccination beliefs and attitudes; weaker beliefs that the vaccination would cause side effects or be unsafe; greater perceived information sufficiency to make an informed decision about COVID-19 vaccination; greater perceived risk of COVID-19 to others; older age; and having been vaccinated for influenza the previous year. Studies conducted before a vaccine was available provided useful data with which to start planning communication strategies about vaccine rollout. With national vaccination programmes currently underway internationally, further research is needed to understand how COVID-19 vaccine acceptance and factors affecting acceptance might have changed now that vaccination has materialised.

Contextual factors such as news stories and media coverage also influence vaccine acceptance [9]. The approval of the COVID-19 vaccines and the rollout of the vaccination programme has been accompanied by considerable press reporting of the differences between the vaccines, including the type of technology used (mRNA vs viral vector [10]), speculation about levels of efficacy observed in clinical trials, and potential variation in effectiveness in a public health context [11]. There was coverage related to two doctors in the UK who had an allergic reaction to the vaccine [12] and some controversy over the deviation from prior clinical trial administration of the required 2 doses of each vaccine 3 weeks apart so that they were administered 12 weeks apart [13], as well as the potential to mix vaccine types [14].

These issues may also have influenced COVID-19 vaccine acceptance. The aim of this study was to investigate associations between COVID-19 vaccination intention and sociodemographic, psychological, and contextual factors in a demographically representative sample of the UK adult population at the start of the COVID-19 vaccination programme rollout.

## METHOD

### Design

We conducted an online cross-sectional survey (13^th^ to 15^th^ January 2021), hosted on Qualtrics.

### Participants

Participants (n=1,500) were recruited through Prolific’s online research panel and were eligible for the study if they were aged eighteen years or over, lived in the UK, and had not completed our previous survey [5] (n>31,000 eligible participants). Quota sampling was used, based on age, sex, and ethnicity, to ensure that the sample was broadly representative of the UK general population. Of 1,508 people who began the survey, 1,503 completed it (99.7% completion rate). Three participants were excluded from the sample as they did not meet quality control checks. Participants were paid £ 2 for a completed survey.

### Measures

Full survey materials are available online [15]. Most items were the same as those in the UK survey reported above [5], which was conducted in July 2020 and consisted of items that were based on previous literature [16-20]. Some further items were added, and some removed or amended to reflect the availability of specific COVID vaccinations and the timing of the survey.

### Personal and clinical characteristics

We asked participants to report their age, gender, ethnicity, religion, highest educational or professional qualifications, current working situation, and total household income. We also asked participants what UK region they lived in, how many people lived in their household, whether they or someone else in their household (if applicable) had a long-standing illness, disability or infirmity and, if so, whether they had received a letter from the NHS recommending that they took extra precautions against coronavirus (‘shielding’) or whether they had a chronic illness that made them clinically vulnerable to serious illness from COVID-19. We also asked whether they or anyone they lived with were classified as obese or were pregnant, and if they worked or volunteered in roles considered critical to the COVID-19 response (‘key worker’ roles).

Lastly, we asked participants whether they had been vaccinated for seasonal influenza last winter and/or had (or intended to be) this winter (yes/no).

### Psychological and contextual factors

Participants were asked to what extent they thought “coronavirus poses a risk to” people in the UK and to themselves personally, on a five-point scale, from “no risk at all” to “major risk”. They were asked if they thought they “have had, or currently have, coronavirus”. Participants could answer “I have definitely had it or definitely have it now”, “I have probably had it or probably have it now”, “I have probably not had it and probably don’t have it now”, and “I have definitely not had it and definitely don’t have it now”. They were also asked if they personally knew anyone who had had COVID-19 (yes/no).

Further, we asked participants a series of eight questions about their attitudes towards COVID-19. They were asked whether, as far as they knew, they were in one of the groups that had so far been offered the vaccine. Participants were then asked if they had been vaccinated (yes, I’ve had one/two doses/no) and if they answered yes, they were asked which vaccine they had received (Pfizer-BioNTech/Oxford University-AstraZeneca). All participants were then asked 21 questions about COVID-19 vaccination. Statements measured theoretical constructs including perceived susceptibility to COVID-19, severity of COVID-19, benefits of a COVID-19 vaccine, barriers to being vaccinated against COVID-19, ability to be vaccinated (self-efficacy), subjective norms, behavioural control, anticipated regret, knowledge, trust in the Government, and trust in the NHS. These items also investigated concerns about commercial profiteering, and participants’ beliefs about vaccination allowing life to get back to ‘normal’ and having to follow social distancing and other restrictions for COVID-19 if vaccinated. Participants rated the statements on an eleven-point scale (0–10) from “strongly disagree” to “strongly agree”. We adjusted the wording to make the grammatical tense either retrospective for those who had received the vaccine or prospective for those who had not. Participants who had not yet received a vaccine were additionally asked how likely they thought it was that they would get side effects from a coronavirus vaccine. We also asked participants if the coronavirus vaccination had been recommended to them by a health care professional and whether their employer did/would want them to have the COVID-19 vaccination. Order of items was quasi-randomized.

### Outcome measure

To measure vaccination intention, we asked participants who had not yet been vaccinated to state how likely they would be to have a COVID-19 vaccination “now that a coronavirus vaccination is available” on an eleven-point scale from “extremely unlikely” (0) to “extremely likely” (10).

We additionally asked participants to report the main reason why they were likely or unlikely to have a coronavirus vaccination in an open-text comment box.

### Ethics

Ethical approval for this study was granted by Keele University’s Research Ethics Committee (reference: PS-200129).

### Sample size

A target sample size of 1500 was chosen to provide a high ratio of cases to estimated parameters in order to avoid overfitting and loss of generalizability in the regression model [21].

### Analysis

To identify variables associated with an intention to have the COVID-19 vaccination in those who had not yet been vaccinated, we constructed a linear regression model. Ordinal and multinomial predictors were converted to dummy variables. To aid interpretation of the model, and to achieve a more parsimonious set of predictor variables, we ran principal component analyses [22] on items investigating beliefs and attitudes about a) COVID-19, and b) COVID-19 vaccination.

Variables entered into the model were selected *a priori* based on their theoretical relevance; no variable selection procedures were employed. Five groups of variables were included in the model: personal and clinical characteristics; seasonal influenza vaccination; general beliefs and attitudes relating to vaccination; beliefs and attitudes relating to COVID-19 illness; and beliefs and attitudes relating to COVID-19 vaccination. The percentage of variance in the outcome variable explained by each predictor was calculated as the squared semipartial correlation for a numerical or binary predictor and the change in *R*^2^ attributable to a set of dummy variables.

As well as fitting the full model, we also added the groups of variables as successive blocks in a hierarchical model, to determine the incremental increase in the adjusted *R*^2^ value as these groups of variables were added to the model.

Due to the large number of predictors in the model, statistical significance was set at *p*≤.01 to control Type 1 errors, and 99% confidence intervals (CIs) were correspondingly calculated for the regression coefficients. Assumptions of the analysis were checked. Analyses were conducted in SPSS 26.

To analyse open-ended responses for reasons why participants were likely or unlikely to have a coronavirus vaccination, we conducted a content analysis using an emergent coding approach, whereby codes were identified from the data rather than *a priori* [23]. Two authors (MC and HD) jointly coded a small sample of statements to understand the scope of the data. They then each independently coded sufficient responses that they achieved a run of 15 statements without encountering any new emerging codes. At this point they compared the codes they had generated and discussed any discrepancies. They then independently applied these codes to the rest of the sample of statements, after which they checked that they had applied the same codes across the statements and discussed and resolved any additional codes and any discrepancies. This process was first applied to those participants who were uncertain about whether they would have the vaccine, then to those who were unlikely to have it, and finally to those participants who were likely to have it.

## RESULTS

Participants were broadly representative of the UK population (mean age 45.6 years, SD=15.6, range 18 to 86; 51% female; 85% white ethnicity; Table 1, see Supplementary Materials 1 for further breakdown). At the time of completing the survey, only 30 respondents had received one or both doses of a coronavirus vaccine.

**Table 1.**
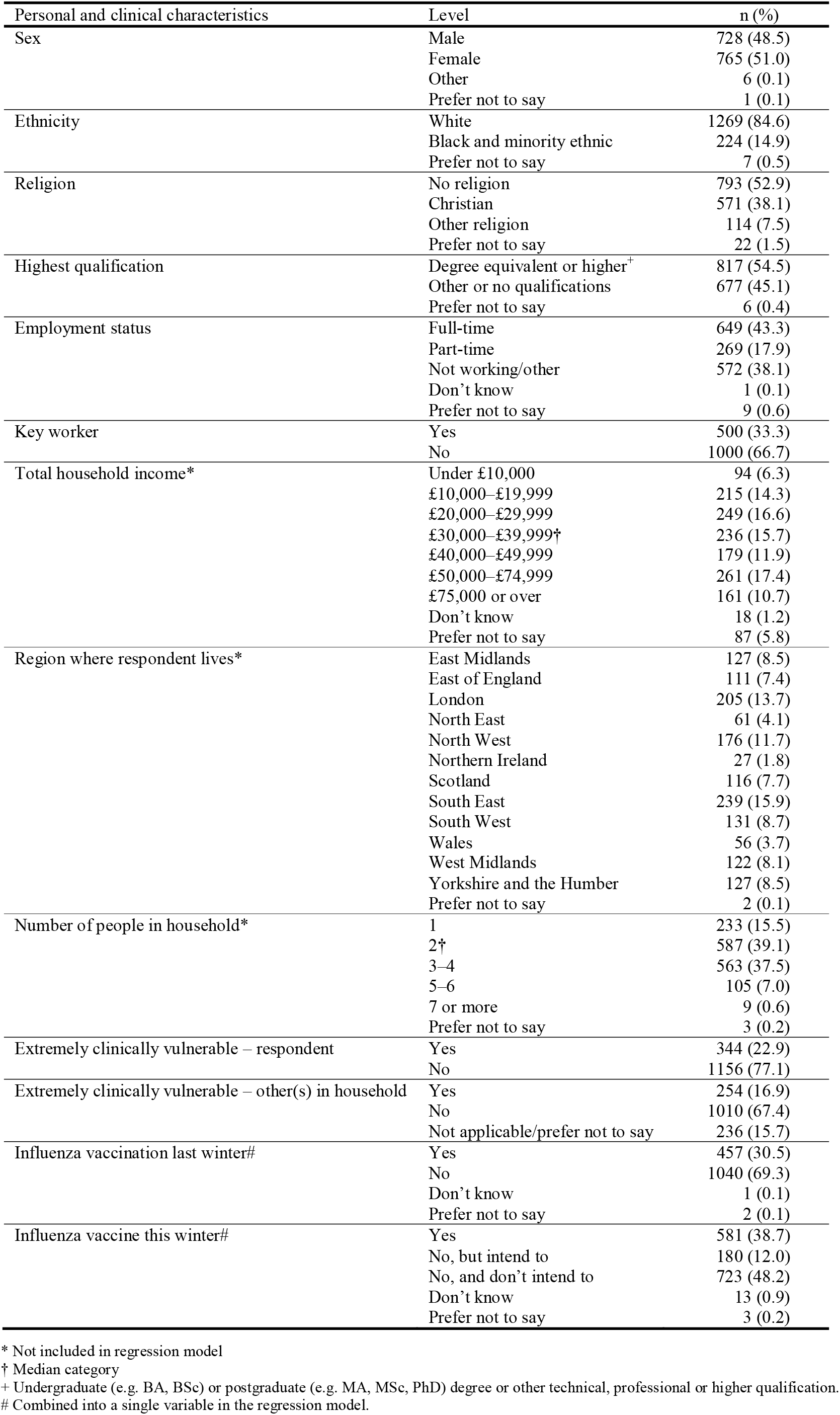
Participant characteristics.

Descriptive statistics for items assessing psychological factors are reported in Tables 2 and 3. These tables show that participants were worried about catching coronavirus and did not believe that it would be a mild illness for them. Approximately three quarters of participants (76.7%) believed COVID-19 posed a moderate or higher risk to them personally. It was also noteworthy that participants reported considerably more trust in the NHS compared to the Government regarding managing the pandemic.

**Table 2.** Descriptive statistics for continuous items measuring beliefs and attitudes about COVID-19 and a COVID-19 vaccination and vaccination intention. Data are mean (standard deviation) on a 0–10 numerical rating scale (0 = strongly disagree, 10 = strongly agree), except where indicated. Also shown is the principal component (as numbered in Table 4) on which the items loaded most, for those items included in the principal components analysis (see Supplementary Materials 2).

|  | Item | Mean (SD) | Related principal component† |
| --- | --- | --- | --- |
| Attitudes and beliefs about COVID-19 | I am worried about catching coronavirus | 6.51 (2.82) | 2 |
|  | I believe that coronavirus would be a mild illness for me | 4.44 (2.73) | 2 |
|  | Too much fuss is being made about the risk of coronavirus* | 2.01 (2.63) | 1 |
|  | We are all responsible for reducing the spread of coronavirus* | 9.23 (1.59) | 1 |
|  | I believe I am immune to coronavirus* | 1.06 (1.91) | 1 |
|  | The coronavirus pandemic has had a big impact on my life | 7.51 (2.33) | 4 |
|  | I trust the NHS to manage the coronavirus pandemic in the UK | 7.39 (2.21) | 3 |
|  | I trust the Government to manage the coronavirus pandemic in the UK | 3.99 (2.98) | 3 |
| Attitudes and beliefs about a COVID-19 vaccination | How likely do you think it is that you would get side effects from a coronavirus vaccination (0 = very unlikely, 10 = very likely) | 4.01 (2.45) | — |
|  | A coronavirus vaccination should be mandatory for everyone who is able to have it | 6.27 (3.60) | 6 |
|  | Without a coronavirus vaccination, I am likely to catch coronavirus | 6.45 (2.44) | 6 |
|  | Two doses of coronavirus vaccination, will protect me against coronavirus | 7.52 (2.22) | 1 |
|  | If I don't get the coronavirus vaccination and end up getting coronavirus, I will regret not getting the vaccination* | 7.92 (3.04) | 6 |
|  | It would be very easy for me to have a coronavirus vaccination* | 7.81 (2.51) | 1 |
|  | The coronavirus vaccination could give me coronavirus | 1.59 (2.17) | 7 |
|  | The way the coronavirus vaccines are being given in the UK goes against the manufacturers' recommendations | 4.89 (2.99) | — |
|  | I might regret getting the coronavirus vaccination if I later experienced side effects from it | 4.42 (3.18) | 7 |
|  | The coronavirus vaccination is too new for me to be confident about getting vaccinated | 4.05 (3.28) | 7 |
|  | Most people will get a coronavirus vaccination | 7.46 (1.75) | 5 |
|  | Other people like me will get a coronavirus vaccination* | 7.94 (2.20) | 5 |
|  | In general, vaccination is a good thing* | 9.02 (1.72) | — |
|  | I am afraid of needles* | 2.77 (3.31) | — |
|  | If I were vaccinated, I think I would not need to follow social distancing and other restrictions for coronavirus | 2.60 (2.82) | 9 |
|  | I know enough about the coronavirus *illness* to make an informed decision about whether or not to get vaccinated | 7.73 (2.45) | 8 |
|  | I know enough about the coronavirus *vaccine* to make an informed decision about whether or not to get vaccinated | 6.79 (2.67) | 8 |
|  | Only people who are at risk of serious illness from coronavirus need to be vaccinated | 2.39 (3.04) | — |
|  | My family would approve of my having a coronavirus vaccination* | 8.58 (2.16) | 5 |
|  | My friends would approve of my having a coronavirus vaccination* | 8.33 (2.09) | 5 |
|  | Widespread coronavirus vaccination is just a way to make money for vaccine manufacturers* | 2.05 (2.62) | — |
|  | The coronavirus vaccine will allow us to get back to 'normal' | 7.24 (2.32) | 5 |
| Vaccination intention | Now that a coronavirus vaccination is available, how likely is it you will have one? (0 = very unlikely, 10 = very likely)* | 8.13 (2.96) | — |

\* Skewed variables; mean values should be interpreted cautiously.
† 1= perceived severity of COVID-19; 2= individual vulnerability to COVID-19; 3= trust in COVID-19 management; 4= impact of COVID-19 on one's life; 5= social norms; 6= the necessity of vaccination; 7= safety of the vaccine; 8= adequacy of information about the vaccine; 9= freedom from restrictions through the vaccine

**Table 3.** Descriptive statistics for categorical and ordinal items measuring beliefs, attitudes and behaviour relating to COVID-19 and a COVID-19 vaccination.

| Item | Level | n (%) |
| --- | --- | --- |
| To what extent do you think coronavirus poses a risk to people in the UK? | No risk at all | 6 (0.4) |
|  | Minor risk | 69 (4.6) |
|  | Moderate risk | 197 (13.1) |
|  | Significant risk | 568 (37.9) |
|  | Major risk | 659 (43.9) |
|  | Don't know | 1 (0.1) |
| To what extent do you think coronavirus poses a risk to you personally? | No risk at all | 37 (2.5) |
|  | Minor risk | 306 (20.4) |
|  | Moderate risk | 539 (35.9) |
|  | Significant risk | 424 (28.3) |
|  | Major risk | 187 (12.5) |
|  | Don't know | 7 (0.5) |
| Do you believe you have had, or currently have, coronavirus? | Definitely not | 460 (30.7) |
|  | Probably not | 621 (41.4) |
|  | Probably | 164 (10.9) |
|  | Definitely | 79 (5.3) |
|  | Don't know | 175 (11.7) |
|  | Prefer not to say | 1 (0.1) |
| Do you personally know anyone (excluding yourself) who has had coronavirus? | Yes | 1153 (76.9) |
|  | No | 336 (22.4) |
|  | Don't know | 10 (0.7) |
|  | Prefer not to say | 1 (0.1) |
| As far as you know, would your employer want you to have the coronavirus vaccination? | Yes | 572 (60.6) |
|  | No | 59 (6.3) |
|  | Don't know | 313 (33.2) |
|  | Not applicable | 556 |

### Principal component analyses

Four components emerged from the principal component analysis on beliefs and attitudes about COVID-19, accounting for 75% of the variance in original items and five components emerged from the principal component analysis investigating items related to a COVID-19 vaccination, accounting for 68% of the variance in the original items (see Supplementary Materials 2)

### Vaccination intention

Participants’ vaccination intention (in participants who had not already received one or both doses) is presented in Figure 1. Vaccination intention exhibited a marked negative skew (mean=8.13, standard deviation=2.96, median=10.00). In order to categorize respondents in terms of their vaccination intention, we applied *a priori* cut-points to the 0–10 scale (with scores of zero to two as “very unlikely”, three to seven as “uncertain” and eight to ten as “very likely”, as per our July 2020 survey [5]). On this basis, 9.3% (95% CI 7.9%, 10.8%) reported being very unlikely to be vaccinated (*n*=136), 17.3% (95% CI 15.4%, 19.3%) reported being uncertain about their likelihood of vaccination (*n*=254), and 73.5% (95% CI 71.2%, 75.7%) reported being very likely to be vaccinated (*n*=1080).

**Figure 1.**
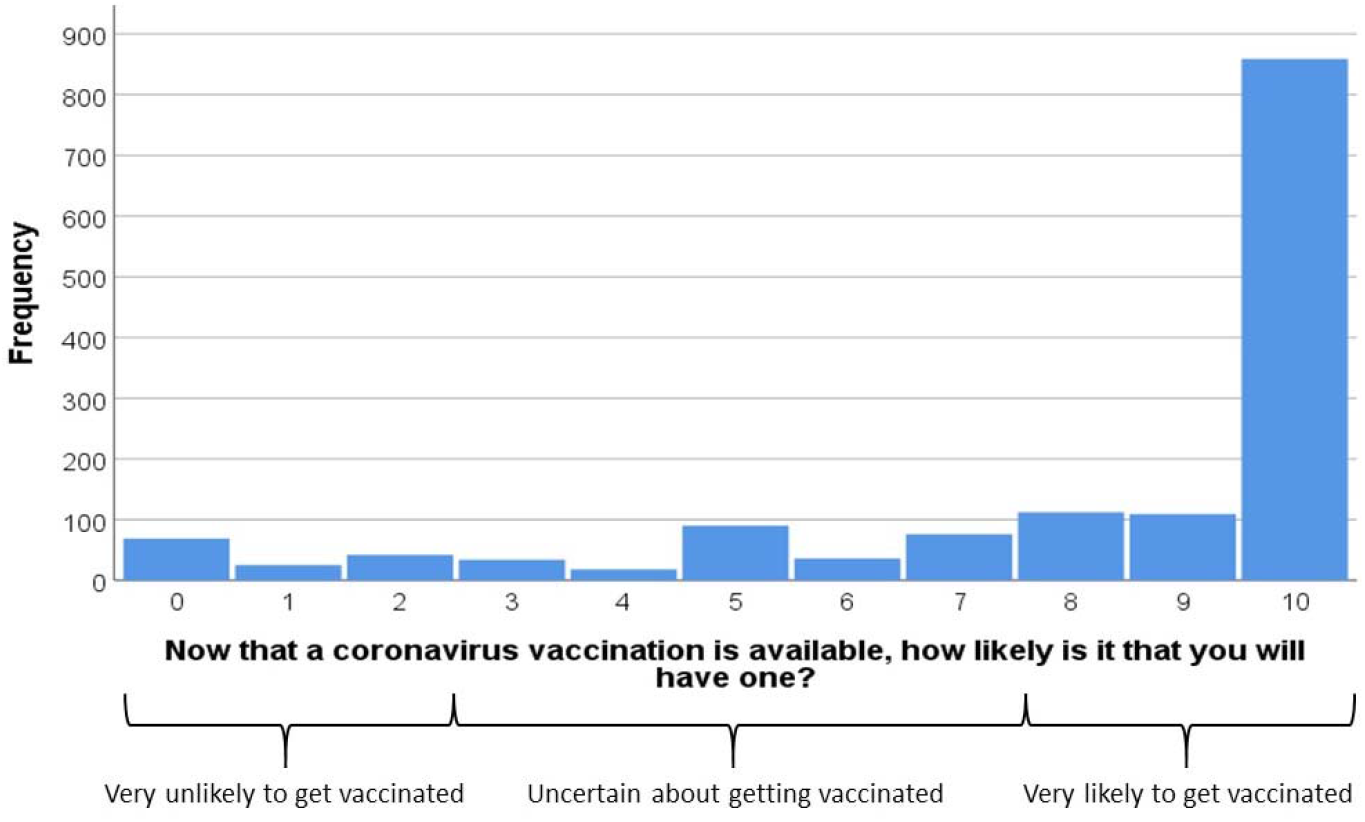
Perceived likelihood of having a vaccination (0=“extremely unlikely” to 10=“extremely likely”). The figure also shows cut-points that we used to categorize respondents in terms of their vaccination intention.

The final model explained 69.8% of the variance in intention to vaccinate (Table 4). Increased likelihood of being vaccinated for COVID-19 was significantly associated with having been vaccinated for influenza last or this winter (or intending to do so this winter), and with all of the components derived from the items relating to COVID-19 vaccination, other than ‘*freedom from restrictions through the vaccine*’. Vaccination intention also showed a significant negative association with beliefs that only people who are at risk of serious illness from coronavirus need to be vaccinated and that widespread coronavirus vaccination is just a way to make money for vaccine manufacturers.

**Table 4.** Results of the full linear regression model analysing associations with vaccination intention (adjusted *R*^2^ = .698). Parameter estimates relate to the full model containing all predictors. The unstandardized regression coefficients represent the change in likelihood of vaccination for a one-unit increase in the predictor variable (or, for dummy variables, a shift from the reference category to the category concerned). The figures under ‘% variance explained’ represent the percentage of variance in the outcome variable uniquely explained by the item (or set of dummy variables) concerned. The model was based on 1401 cases with complete data.

| Predictor variable | Level | Standardized coefficient | Unstandardized coefficient | 99% confidence interval | p value | % variance explained |
| --- | --- | --- | --- | --- | --- | --- |
| Block 1 – personal and clinical characteristics |  |  |  |  |  |  |
| Age | Years | .046 | .008 | –.001, .018 | .023 | .11 |
| Sex (reference: female) | Male | –.024 | –.135 | –.364, .094 | .129 | .05 |
| Ethnicity (reference: black and minority ethnic) | White | –.002 | –.016 | –.378, .346 | .907 | <.01 |
| Religion (reference: none) |  |  |  |  | .935 | <.01 |
|  | Christian | .000 | .003 | –.249, .255 |  |  |
|  | Other | .006 | .068 | –.414, .549 |  |  |
| Qualifications (reference: other) | Degree equivalent or higher | .028 | .162 | –.070, .393 | .066 | .07 |
| Employment status (reference: not working/other) |  |  |  |  | .336 | .05 |
|  | Part-time | .014 | .105 | –.236, .446 |  |  |
|  | Full-time | –.014 | –.080 | –.356, .197 |  |  |
| Key worker (reference: not key worker) | Key worker | –.003 | –.018 | –.280, .245 | .863 | <.01 |
| Extremely clinically vulnerable – self (reference: no) | Yes | –.023 | –.155 | –.444, .134 | .166 | .04 |
| Extremely clinically vulnerable – household member (reference: no) | Yes | –.022 | –.173 | –.474, .129 | .140 | .05 |
| Block 2 – previous influenza vaccination |  |  |  |  |  |  |
| Did you/will you have a vaccination for influenza last/this winter? (reference: no) | Yes | .047 | .270 | .012, .528 | .007* | .14 |
| Block 3 – general vaccination beliefs and attitudes |  |  |  |  |  |  |
| Vaccination is generally good (0–10) | 0–10 scale | .011 | .018 | –.079, .116 | .627 | .01 |
| I am afraid of needles (0–10) | 0–10 scale | .016 | .014 | –.021, .049 | .295 | .02 |
| Block 4 – beliefs and attitudes about COVID-19 |  |  |  |  |  |  |
| Perceived risk of COVID-19 to people in the UK (reference: major) |  |  |  |  | .822 | .02 |
|  | None/minor | –.012 | –.166 | –.897, .565 |  |  |
|  | Moderate | .007 | .063 | –.380, .507 |  |  |
|  | Significant | .002 | .015 | –.261, .290 |  |  |
| Perceived risk of COVID-19 to oneself (reference: major) |  |  |  |  | .021 | .21 |
|  | None/minor | .059 | .407 | –.190, 1.005 |  |  |
|  | Moderate | .082 | .493 | .036, .951 |  |  |
|  | Significant | .067 | .429 | .034, .824 |  |  |
| Do you have/have you had COVID-19? (reference: probably/definitely) |  |  |  |  | .288 | .08 |
|  | Probably not | .028 | .163 | –.180, .586 |  |  |
|  | Definitely not | –.002 | –.010 | –.375, .355 |  |  |
|  | Don’t know | .018 | .164 | –.283, .610 |  |  |
| Do you know anybody who has had COVID-19? (reference: no) | Yes | .005 | .032 | –.246, .310 | .764 | .02 |
| Component 1: perceived severity of COVID-19 |  | –.028 | –.083 | –.257, .091 | .219 | .03 |
| Component 2: individual vulnerability to COVID-19 |  | –.045 | –.130 | –.308, .048 | .060 | .08 |
| Component 3: trust in COVID-19 management |  | –.005 | –.015 | –.144, .114 | .766 | <.01 |
| Component 4: impact of COVID-19 on one’s life |  | –.011 | –.031 | –.156, .093 | .513 | .01 |
| Block 5 – beliefs and attitudes about COVID-19 vaccination |  |  |  |  |  |  |
| Component 5: social norms |  | .378 | 1.106 | .954, 1.259 | <.001* | 7.55 |
| Component 6: the necessity of vaccination |  | .460 | 1.329 | 1.172, 1.487 | <.001* | 10.20 |
| Component 7: safety of the vaccine |  | .370 | 1.068 | .923, 1.212 | <.001* | 7.83 |
| Component 8: adequacy of information about the vaccine |  | .148 | .431 | .308, .554 | <.001* | 1.76 |
| Component 9: freedom from restrictions through the vaccine |  | .015 | .043 | -.077, .164 | .353 | .02 |
| The way the coronavirus vaccines are being given in the UK goes against the manufacturers' recommendations |  | .005 | .005 | -.035, .045 | .751 | <.01 |
| Only people who are at risk of serious illness from coronavirus need to be vaccinated | 0–10 scale | -.064 | -.061 | -.106, -.016 | .001* | .26 |
| Widespread coronavirus vaccination is just a way to make money for vaccine manufacturers | 0–10 scale | -.060 | -.067 | -.128, -.006 | .004* | .18 |

\* $p \leq .01$

The principal component that related to the necessity of vaccination explained more variance in vaccination intention than any other predictor in the statistical model, followed by the principal components concerning social acceptance and safety of the vaccine. Other significant predictors only explained small percentages of variance.

When the groups of variables were entered hierarchically as blocks, we could infer the percentage of additional variance explained by each block from the change in incremental adjusted *R*^2^. Personal and clinical characteristics (block 1) alone explained very little (8.8%) of the variance in intention to be vaccinated. When previous influenza vaccination (block 2) was added, it explained an additional 6.4% of the variance. Adding general vaccination beliefs and attitudes (block 3) resulted in the largest increase in proportion (25.1%) of explained variance (though in the full model the predictors in this group were no longer significant). When beliefs and attitudes about COVID-19 (block 4) were added to the model, they explained 6.5% more of the variance in vaccination intention. Adding positive beliefs and attitudes about a COVID-19 vaccination (block 5) explained a further 23.0% of the variance. Each block explained a statistically significant proportion of the variance (p<.001 in each case).

## Content Analysis

Of the 1470 participants who had not yet received a vaccine and were asked to give a reason for the score provided on the likelihood of having the vaccination question, 1461 participants (99.4%) provided a response. Answers ranged from one word to 455 words (mean=20.3, SD=20.6). The content analysis generated 102 unique codes. The codes were then further organised into themes and these, along with a frequency count of comments per theme, are presented in Table 5. A breakdown of codes and themes is provided in Supplementary Materials 3.

**Table 5.** Thematic categorization of codes generated by content analysis of reasons for likelihood of having, or not having, the COVID-19 vaccination, including breakdown by likelihood of having the vaccination (likely, uncertain, likely).

| Theme name | Illustrative code | Number of comments per theme (likely, uncertain, unlikely) |
| --- | --- | --- |
| Self-protection (including health reasons) | Perceived high personal risk of disease severity | 675 (651, 22, 2) |
| To protect others | Protecting the wider community | 667 (609, 54, 4) |
| To end the pandemic and its negative impacts | To end lockdown | 345 (331, 14, 0) |
| Confidence in vaccine and authority | The vaccine is effective | 317 (276, 39, 2) |
| Safety concerns about the COVID-19 vaccine | Concerns re quick development of the vaccine | 226 (28, 110, 88) |
| Concerns re details of vaccine (other than safety) | Concerns re dose time scale | 173 (22, 89, 62) |
| Low risk/no personal need for vaccine | Only high-risk groups need the vaccine | 144 (25, 75, 44) |
| Concern about health effects of COVID-19 | Concerns re long term side effects of virus | 77 (71, 6, 0) |
| Other (miscellaneous) | Currently no access due to visa status | 67 (46, 13, 8) |
| Precontemplation/not made decision | Not offered yet | 64 (12, 44, 8) |
| Unspecified concerns about COVID-19 vaccine | Anxiety re the vaccine | 62 (0, 24, 38) |
| To travel/move around more freely | Wanting to visit family | 55 (42, 13, 0) |
| Specific health concerns about the COVID-19 vaccine | Fertility concerns | 39 (5, 19, 15) |
| Avoid/delay having the vaccine by waiting | Wanting others to test it first | 36 (5, 25, 6) |
| Lack of trust in authority | Lack of trust in media transparency | 33 (7, 7, 19) |
| General vaccine concerns | Fear of needles | 30 (4, 8, 18) |

The two most frequently cited reasons to support the score participants gave on the likelihood question related to protecting themselves or others. These were primarily reasons given by participants who indicated they were likely to have the vaccine. In comparison, the most frequently provided reasons provided by participants who were uncertain or unlikely to have the vaccine were related to safety concerns about the COVID-19 vaccine.

## DISCUSSION

The UK government has set a target of offering a first dose of a COVID-19 vaccine to all adults in the UK by the end of July 2021 [24]. In this study conducted in January 2021 three-quarters of participants reported being very likely to have the vaccination. This is higher than the 64% who reported being very likely to have the vaccination in our study conducted in July 2020 [5] and is consistent with increases in vaccination intention reported elsewhere. For example, in a recent (March 2021) YouGov poll, 86% of respondents had either already been vaccinated or reported that they would get the vaccine [25]. Despite the relatively high intention reported in our study and recent polling, we cannot be complacent about uptake. News stories are emerging every week about different variants of the virus as well as issues around differential uptake of individual vaccines across the world [e.g. 26] and it is important to understand the factors associated with intention in order to maximize uptake and offset any adverse media reporting.

Our results indicate that greater intention to have the COVID-19 vaccination was associated with having been vaccinated for influenza last or this winter or intending to be this winter. This is consistent with our previous findings [5] as well as with findings from the US [27] and Europe [28]. However, several of the COVID-19 vaccination beliefs and attitudes explained a more substantial proportion of the variance in vaccination intention. Intention was associated with greater perceived social norms around COVID-19 vaccination and greater perceived necessity of vaccination. These items map onto theoretical constructs that have previously been shown to influence uptake of health behaviours: subjective norms and perceived susceptibility [5]. Previous studies exploring vaccine intentions have also found high levels of positive social norms favouring the vaccine [29] and that intention is associated with increased levels of concern related to the risks of the disease [28, 30]. We also found that lower intention was associated with reduced belief in the safety of the vaccine and this has been found consistently across studies exploring COVID-19 vaccine intentions [7, 8] and vaccine hesitancy [8]. This was also reflected in the content analysis of participants’ open-ended responses, in which issues related to vaccine safety were the most frequently identified reason for lower vaccination intention in the participants we classified as uncertain or very unlikely to have the vaccine. This is also consistent with the free text responses given in an English study exploring vaccine acceptability conducted April to May 2020 [7]. Since there was less than a year between the genetic code of COVID-19 being made public and the first COVID-19 vaccine being approved, this belief is perhaps unsurprising. However, it does make it essential that sufficient engagement with the public’s concerns takes place and good-quality and credible information continues to be made available about the vaccine. This recommendation is reinforced by the association of perceived adequacy of information about the vaccine to facilitate an informed decision and vaccination intention in our survey. Since free-text responses related to safety were the most frequent category of response from uncertain responders, it is likely that any reporting of safety concerns in the media may well shift the balance in favour of not being vaccinated, as has been observed previously [31]. It is imperative that halting the rollout of the vaccination programme because of unproven risks, as seen in some European countries in March 2021 [26], should be avoided, as this is likely to damage uptake once vaccination is restarted.

Vaccination intention in our study was also associated with a weaker belief that only people who are at risk of serious illness from coronavirus need to be vaccinated and free-text responses associated with protecting others were frequently given to explain intention to have the vaccine. Statements about protecting others were only second in the content analysis to those relating to self-protection, such as a perceived high personal risk of disease severity and this is consistent with previous research on COVID-19 vaccine intentions [7].

Several previous studies have found that various socio-demographic factors are associated with COVID-19 vaccine intention such as age [5, 30], gender [28, 32] and ethnicity [7, 32]. We did not find this in our study and it is not entirely clear why that might be. The lack of impact of ethnicity on intention is perhaps the most striking absence, given both previous research and evidence from actual uptake in the UK, which shows that a significantly smaller proportion of ethnic minority compared to white health care workers have had the COVID-19 vaccine [33]. We recruited a demographically representative sample based on age, gender and ethnicity. However, the relatively low number of participants from ethnic minority backgrounds necessitated collapsing our data across these categories, which may explain why ethnicity was not associated with intention in our study. The lack of an association with age may reflect the fact that the government plans to roll out the vaccination to all adults across the UK. Studies using purposive sampling techniques are required to capture and quantify uptake and associated attitudinal differences across different population cohorts.

Finally, intention to have the COVID-19 vaccine was associated with a weaker belief that widespread coronavirus vaccination is just a way to make money for vaccine manufacturers. This is consistent with research from Hong Kong [34] in which higher levels of trust in the vaccine manufacturer were associated with increased willingness to have the COVID-19 vaccine.

Limitations to this study include that we measured self-reported intention rather than actual uptake. Intention is usually higher than uptake; however, vaccine intention predicts vaccine uptake and so acts as a useful proxy in the early stages of vaccination rollout. Also, the survey is cross-sectional, so we are unable to infer causality between attitudinal factors and intention.

To our knowledge, this is the first peer-reviewed study investigating intention to receive a COVID-19 vaccination in a demographically representative sample of the UK population since the COVID-19 vaccination rollout began in December 2020. Three-quarters of our sample reported being very likely to have the COVID-19 vaccination. However, since vaccine uptake may well be lower than vaccine intention, it is important to understand the factors associated with intention and to ensure that communication and engagement strategies related to the vaccination are informed by those factors. Going forward, it is not yet known for how long COVID-19 vaccines confer immunity or how effective they will continue to be against emerging strains as the virus mutates and, consequently, whether booster vaccines may be required [24]. In order to ensure the success of the current vaccination rollout and any subsequent vaccination waves, our findings underline the importance of ongoing clear communication informed by theoretical constructs related to COVID-19 vaccination beliefs and attitudes, and the need for such communication to emphasize social acceptance of the vaccination, the importance of vaccination to stop the spread of COVID-19, even in the absence of underlying risk factors, and the safety of the vaccination.

## Data Availability

Data are available through the Open Science Framework (OSF) site. A peer review link to the data is here: https://osf.io/ewch3/?view_only=8d25cf3247e240e28e61c9b8f5d04f01. On acceptance the data will be fully open.

## TRANSPARENCY DECLARATION

The authors affirm that the manuscript is an honest, accurate, and transparent account of the study being reported; that no important aspects of the study have been omitted; and that any discrepancies from the study as originally planned have been explained.

## DATA SHARING STATEMENT

Data are available online [15].

## SUPPLEMENTARY MATERIAL

**Supplementary Table 1.**
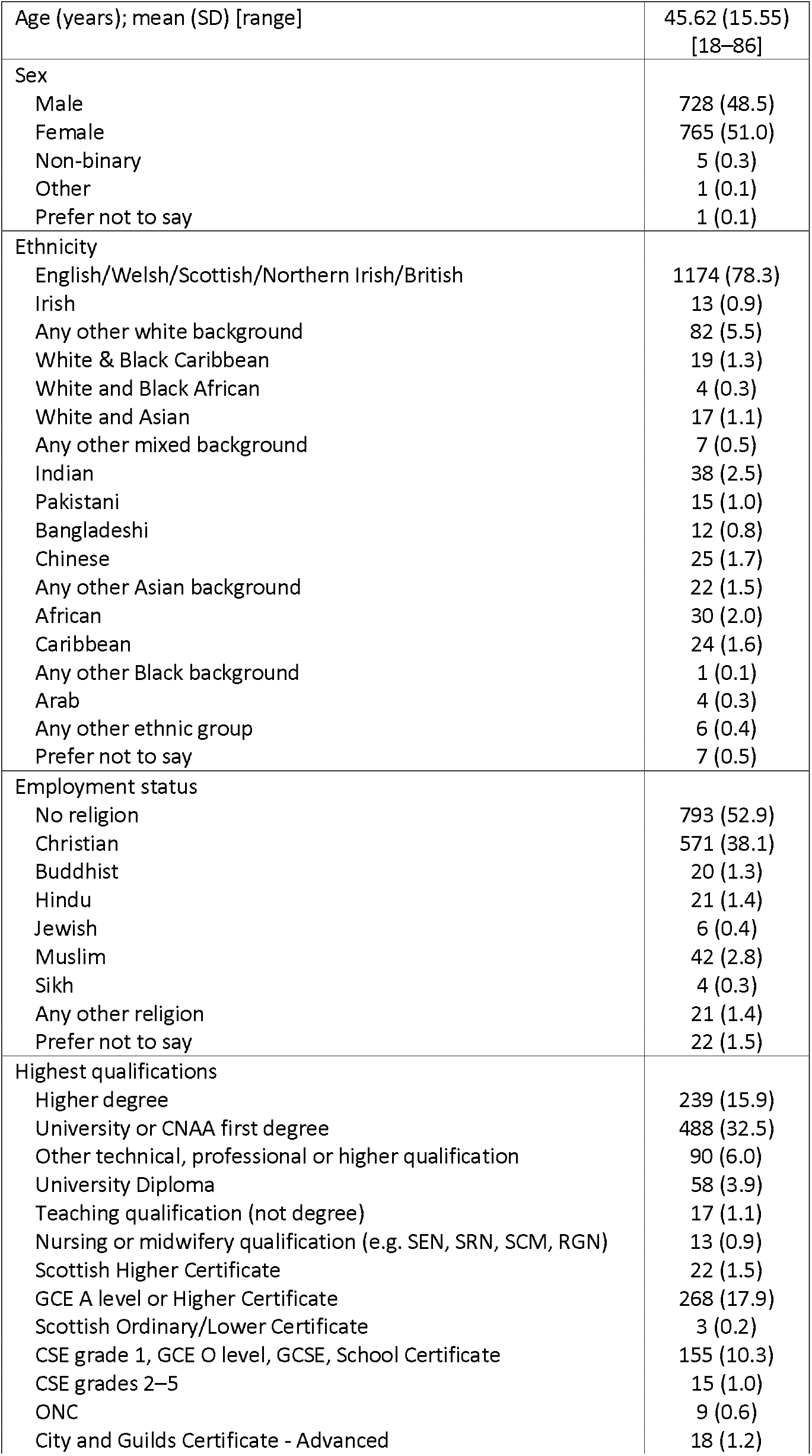
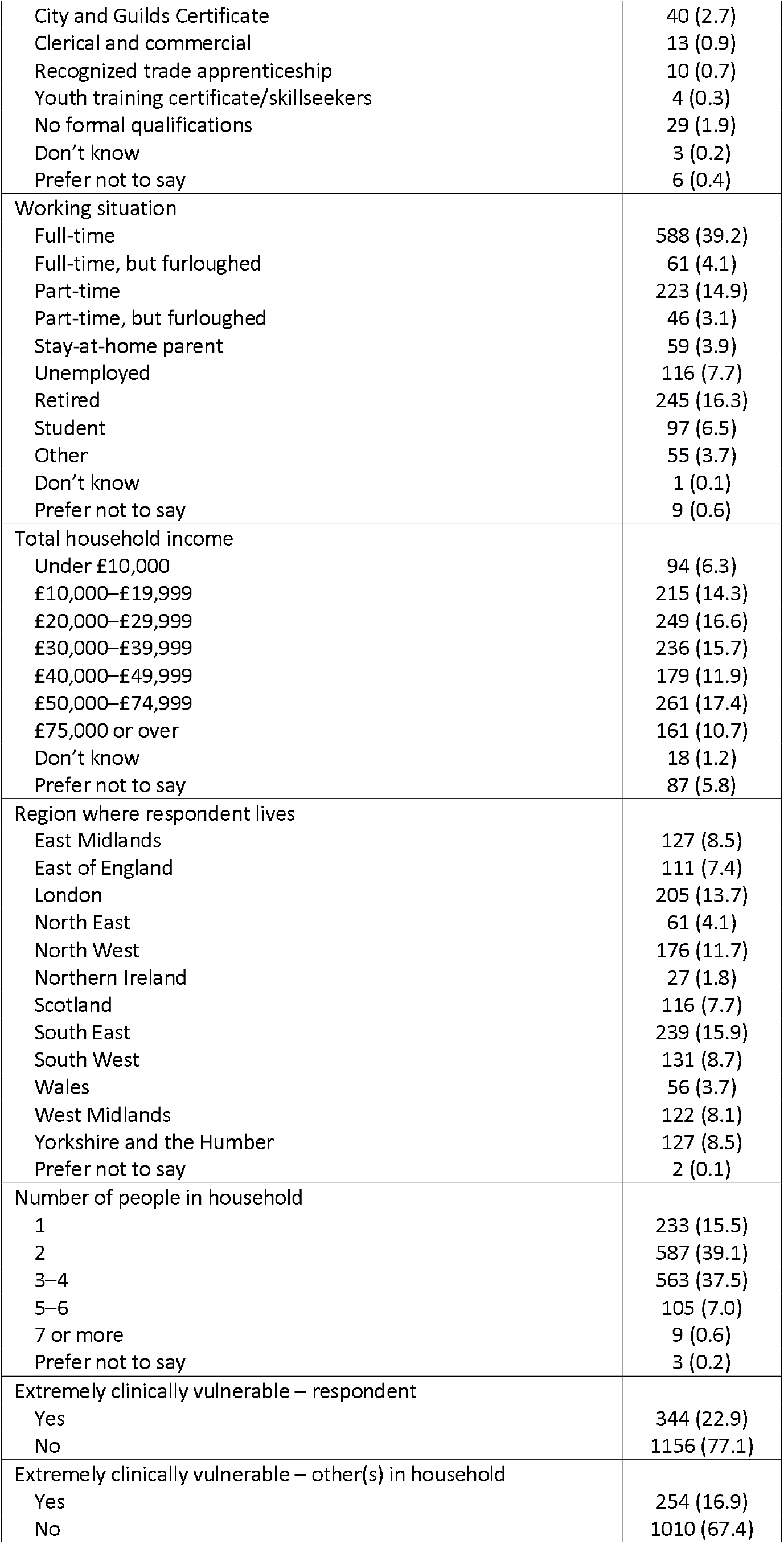
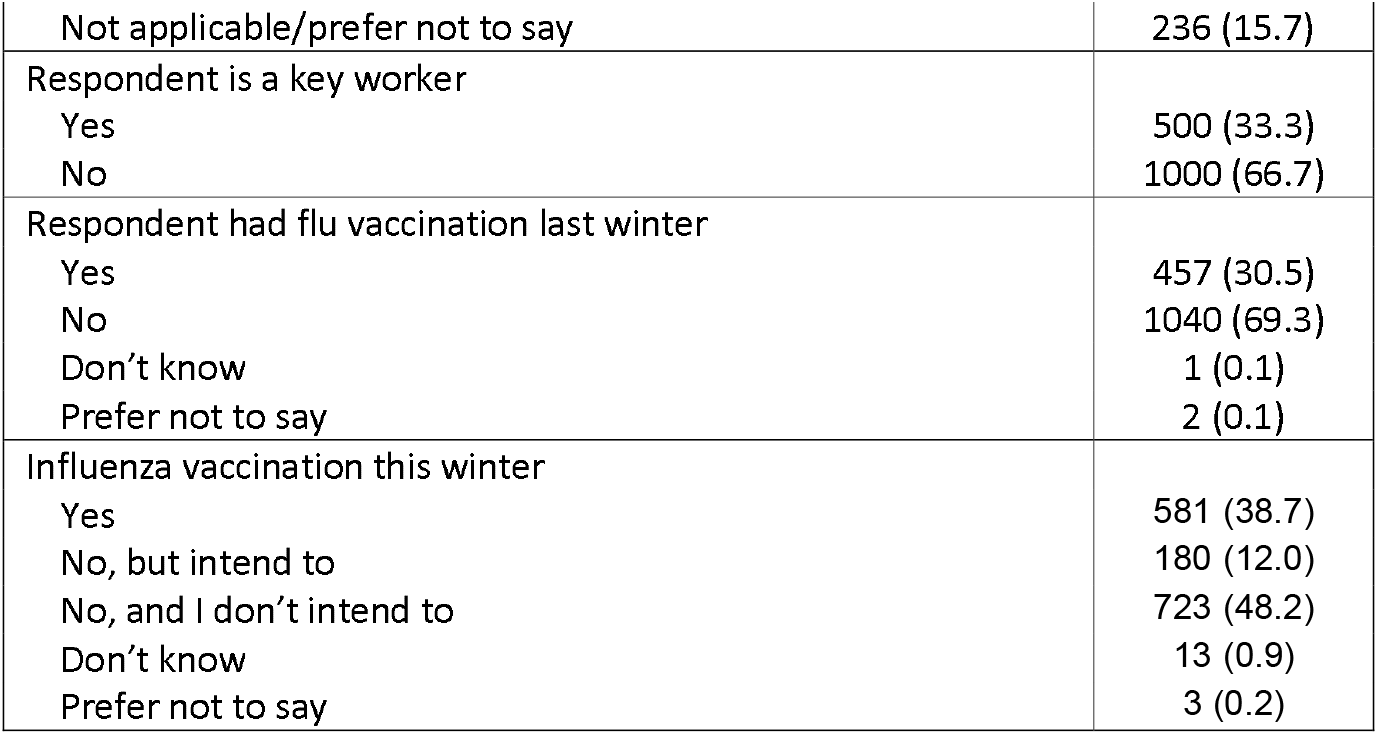
Respondent characteristics. Data are frequencies (%) except where indicated.

**Supplementary Table 2:** Principal components analyses for COVID-19 and COVID-19 vaccination items

**Rotated Component Matrix<sup>a</sup>**
|  | Component |  |  |  |
| --- | --- | --- | --- | --- |
|  | 1 | 2 | 3 | 4 |
| I am worried about catching coronavirus | -.446 | -.671 |  |  |
| I believe that coronavirus would be a mild illness for me |  | .898 |  |  |
| Too much fuss is being made about the risk of coronavirus | .733 |  |  |  |
| We are all responsible for reducing the spread of the coronavirus | -.731 |  |  |  |
| I believe I am immune to coronavirus | .728 |  |  |  |
| The coronavirus pandemic has had a big impact on my life |  |  |  | .972 |
| I trust the NHS to manage the coronavirus pandemic in the UK | -.449 |  | .698 |  |
| I trust the Government to manage the coronavirus pandemic in the UK |  |  | .885 |  |

**Rotated Component Matrix<sup>a</sup>**
|  | Component |  |  |  |  |
| --- | --- | --- | --- | --- | --- |
|  | 5 | 6 | 7 | 8 | 9 |
| Coronavirus vaccination should be mandatory for everyone who is able to have it |  | .718 |  |  |  |
| Without a coronavirus vaccination, I am likely to catch coronavirus |  | .765 |  |  |  |
| Two doses of coronavirus vaccination will protect me against coronavirus | .467 | .445 | .407 |  |  |
| If I don't get the coronavirus vaccination and end up getting coronavirus, I will regret not getting the vaccination |  | .599 |  |  |  |
| It would be easy for me to have the coronavirus vaccination | .440 |  |  |  |  |
| The coronavirus vaccination could give me coronavirus |  |  | -.715 |  |  |
| I might regret getting the coronavirus vaccination if I later experienced side effects from it |  |  | -.703 |  |  |
| The coronavirus vaccination is too new for me to be confident about it |  |  | -.698 |  |  |
| Most people will get the coronavirus vaccination | .780 |  |  |  |  |
| Other people like me will get the coronavirus vaccination | .690 |  |  |  |  |
| If I were vaccinated, I think I would not need to follow social distancing and other restrictions for coronavirus |  |  |  |  | .928 |
| I know enough about the coronavirus illness to make an informed decision about whether or not to get vaccinated |  |  |  | .863 |  |
| I know enough about the coronavirus vaccine to make an informed decision about whether or not to get vaccinated |  |  |  | .805 |  |
| My family would approve of my having the coronavirus vaccination | .606 | .410 |  |  |  |
| My friends would approve of my having the coronavirus vaccination | .625 |  |  |  |  |
| The coronavirus vaccine will allow us to get back to 'normal' | .525 |  |  |  |  |

Extraction Method: Principal Component Analysis.
Rotation Method: Varimax with Kaiser Normalization.<sup>a</sup>
a. Rotation converged in 7 iterations.

**Supplementary Table 3:** Content analysis codes and themes.

| Themes and codes | Likely | Uncertain | Unlikely | Total | % of ppts |
| --- | --- | --- | --- | --- | --- |
| **SELF PROTECTION (incl health reasons)** |  |  |  |  |  |
| Protecting self | 510 | 20 | 0 | 530 | 35.33 |
| High-risk category | 52 | 2 | 2 | 56 | 3.73 |
| Vulnerable due to underlying health condition | 54 | 0 | 0 | 54 | 3.60 |
| Perceived high personal risk of disease severity | 20 | 0 | 0 | 20 | 1.33 |
| Negative experience with COVID in the past | 13 | 0 | 0 | 13 | 0.87 |
| Following medical advice | 2 | 0 | 0 | 2 | 0.13 |
|  | **651** | **22** | **2** | **675** | **45.00** |
| **PROTECT OTHERS** |  |  |  |  |  |
| Protecting the wider community | 220 | 27 | 1 | 248 | 16.53 |
| Protecting family | 91 | 9 | 0 | 100 | 6.67 |
| Vaccine is a civic duty/social responsibility | 83 | 0 | 0 | 83 | 5.53 |
| To reduce the spread | 84 | 0 | 0 | 84 | 5.60 |
| Mass vaccination needed | 63 | 0 | 0 | 63 | 4.20 |
| To gain herd immunity | 31 | 0 | 0 | 31 | 2.07 |
| Altruism | 11 | 14 | 3 | 28 | 1.87 |
| Protecting the vulnerable | 18 | 4 | 0 | 22 | 1.47 |
| Protecting those that cannot have the vaccine | 8 | 0 | 0 | 8 | 0.53 |
|  | **609** | **54** | **4** | **667** | **44.47** |
| **TO END PANDEMIC AND ITS NEGATIVE IMPACTS** |  |  |  |  |  |
| Regain normality | 204 | 14 | 0 | 218 | 14.53 |
| To overcome the pandemic | 88 | 0 | 0 | 88 | 5.87 |
| Health system relief | 15 | 0 | 0 | 15 | 1.00 |
| To end lockdown | 12 | 0 | 0 | 12 | 0.80 |
| To alleviate mental health strain | 5 | 0 | 0 | 5 | 0.33 |
| To ease the economy | 3 | 0 | 0 | 3 | 0.20 |
| Wanting to find work/resume working | 3 | 0 | 0 | 3 | 0.20 |
| Impact of lockdown on physical health | 1 | 0 | 0 | 1 | 0.07 |
|  | **331** | **14** | **0** | **345** | **23.00** |
| **CONFIDENCE IN VACCINE AND AUTHORITY** |  |  |  |  |  |
| Would have it if offered/asked | 78 | 34 | 1 | 113 | 7.53 |
| Pro-vaccine in general | 44 | 0 | 0 | 44 | 2.93 |
| No safety concerns re the vaccine | 32 | 3 | 1 | 36 | 2.40 |
| Not reason not to have vaccine | 29 | 0 | 0 | 29 | 1.93 |
| Trust in science | 28 | 0 | 0 | 28 | 1.87 |
| The vaccine is effective | 16 | 0 | 0 | 16 | 1.07 |
| Preferring to have the vaccine | 16 | 0 | 0 | 16 | 1.07 |
| Vaccine is available | 12 | 0 | 0 | 12 | 0.80 |
| Trust in vaccine | 8 | 0 | 0 | 8 | 0.53 |
| Trust in government | 5 | 0 | 0 | 5 | 0.33 |
| Vaccine should be mandatory | 4 | 0 | 0 | 4 | 0.27 |
| Would have if free to access | 2 | 2 | 0 | 4 | 0.27 |
| Trust in NHS | 2 | 0 | 0 | 2 | 0.13 |
|  | **276** | **39** | **2** | **317** | **21.13** |
| **SAFETY CONCERNS RE COVID VACCINE** |  |  |  |  |  |
| Concerns re quick development of the vaccine | 3 | 25 | 33 | 61 | 4.07 |
| Concerns re long term side effects of vaccine | 13 | 27 | 20 | 60 | 4.00 |
| Concerns re vaccine side effects | 10 | 33 | 17 | 60 | 4.00 |
| Concerns re vaccine safety | 2 | 24 | 13 | 39 | 2.60 |
| Vaccine creates a new strand | 0 | 0 | 1 | 1 | 0.07 |
| Flu jab increases vulnerability to COVID and vice versa | 0 | 1 | 0 | 1 | 0.07 |
| Vaccine more risky than virus | 0 | 0 | 4 | 4 | 0.27 |
|  | **28** | **110** | **88** | **226** | **15.07** |
| **CONCERNS RE DETAILS OF VACCINE (OTHER THAN SAFETY)** |  |  |  |  |  |
| Lack of research re the vaccine | 7 | 32 | 34 | 73 | 4.87 |
| Concerns re effectiveness of vaccine | 3 | 22 | 11 | 36 | 2.40 |
| Vaccine does not stop COVID transmission | 1 | 8 | 4 | 13 | 0.87 |
| Concerns re vaccine composition | 0 | 7 | 6 | 13 | 0.87 |
| Concerns re dose time scale | 4 | 5 | 3 | 12 | 0.80 |
| Lack of knowledge about vaccine | 1 | 7 | 1 | 9 | 0.60 |
| Doubt re effectiveness of vaccine against different variants | 1 | 4 | 0 | 5 | 0.33 |
| Concern re number of doses | 0 | 0 | 3 | 3 | 0.20 |
| Concerns re vaccine mix and match | 1 | 2 | 0 | 3 | 0.20 |
| Unsure which vaccine | 1 | 2 | 0 | 3 | 0.20 |
| Wanting a specific vaccine | 3 | 0 | 0 | 3 | 0.20 |
|  | **22** | **89** | **62** | **173** | **11.53** |
| **LOW RISK/NO PERSONAL NEED FOR VACCINE** |  |  |  |  |  |
| Low-risk category | 16 | 47 | 16 | 79 | 5.27 |
| Perceived low personal risk of disease severity | 5 | 11 | 12 | 28 | 1.87 |
| No personal need for the vaccine | 0 | 9 | 10 | 19 | 1.27 |
| Others need it more | 4 | 7 | 2 | 13 | 0.87 |
| Only high-risk groups need the vaccine | 0 | 1 | 4 | 5 | 0.33 |
|  | **25** | **75** | **44** | **144** | **9.60** |
| **CONCERN RE HEALTH EFFECTS OF VIRUS** |  |  |  |  |  |
| Vaccine reduces disease severity/fatality | 30 | 0 | 0 | 30 | 2.00 |
| Virus more risky than vaccine | 18 | 0 | 0 | 18 | 1.20 |
| Concerns re long term side effects of virus | 9 | 6 | 0 | 15 | 1.00 |
| Anxiety re the virus | 12 | 0 | 0 | 12 | 0.80 |
| Immunity after infection doesn't last long | 1 | 0 | 0 | 1 | 0.07 |
| No treatments available | 1 | 0 | 0 | 1 | 0.07 |
|  | **71** | **6** | **0** | **77** | **5.13** |
| **OTHER** |  |  |  |  |  |
| Work requirement | 35 | 6 | 0 | 41 | 2.73 |
| Likely already had COVID | 9 | 4 | 8 | 21 | 1.40 |
| Social influence | 1 | 1 | 0 | 2 | 0.13 |
| Unable to travel to access the vaccine | 1 | 1 | 0 | 2 | 0.13 |
| Currently no access due to visa status | 0 | 1 | 0 | 1 | 0.07 |
|  | **46** | **13** | **8** | **67** | **4.47** |
| **PRECONTEMPLATION/DECISION** |  |  |  |  |  |
| Not offered yet | 11 | 18 | 6 | 35 | 2.33 |
| Undecided | 1 | 24 | 2 | 27 | 1.80 |
| Not yet contemplated | 0 | 2 | 0 | 2 | 0.13 |
|  | **12** | **44** | **8** | **64** | **4.27** |
| **UNSPECIFIED CONCERNS RE VACCINE** |  |  |  |  |  |
| Lack of trust in the vaccine | 0 | 7 | 20 | 27 | 1.80 |
| Preferring not to have the vaccine | 0 | 6 | 10 | 16 | 1.07 |
| Other preventative measures are more effective | 0 | 6 | 1 | 7 | 0.47 |
| Conflicting information | 0 | 2 | 4 | 6 | 0.40 |
| Anxiety re the vaccine | 0 | 3 | 1 | 4 | 0.27 |
| Lack of liability from manufacturers | 0 | 0 | 2 | 2 | 0.13 |
|  | **0** | **24** | **38** | **62** | **4.13** |
| **TO TRAVEL/MOVE AROUND FREELY** |  |  |  |  |  |
| Wanting to travel | 17 | 8 | 0 | 25 | 1.67 |
| Wanting to visit family | 16 | 1 | 0 | 17 | 1.13 |
| For personal freedom | 4 | 4 | 0 | 8 | 0.53 |
| Wanting to visit friends | 4 | 0 | 0 | 4 | 0.27 |
| To gain an immunity passport | 1 | 0 | 0 | 1 | 0.07 |
|  | **42** | **13** | **0** | **55** | **3.67** |
| **SPECIFIC HEALTH CONCERNS RE VACCINE** |  |  |  |  |  |
| Pregnancy concerns | 1 | 5 | 8 | 14 | 0.93 |
| Allergy concerns | 2 | 5 | 3 | 10 | 0.67 |
| Fertility concerns | 1 | 6 | 2 | 9 | 0.60 |
| Interference of the vaccine with other health conditions | 1 | 3 | 1 | 5 | 0.33 |
| Perceived high risk due to mental health | 0 | 0 | 1 | 1 | 0.07 |
|  | **5** | **19** | **15** | **39** | **2.60** |
| **AVOID/DELAY BY WAITING** |  |  |  |  |  |
| Wanting others to test it first | 0 | 16 | 2 | 18 | 1.20 |
| If vaccine becomes a requirement | 5 | 6 | 2 | 13 | 0.87 |
| Hoping for herd immunity to avoid vaccination | 0 | 3 | 2 | 5 | 0.33 |
|  | **5** | **25** | **6** | **36** | **2.40** |
| **LACK OF TRUST IN AUTHORITY** |  |  |  |  |  |
| Lack of trust in government | 6 | 4 | 11 | 21 | 1.40 |
| Lack of trust in science | 1 | 0 | 5 | 6 | 0.40 |
| Lack of trust in media transparency | 0 | 0 | 3 | 3 | 0.20 |
| People in power's influence | 0 | 2 | 0 | 2 | 0.13 |
| Vaccine too politicised | 0 | 1 | 0 | 1 | 0.07 |
|  | **7** | **7** | **19** | **33** | **2.20** |
| **GENERAL VACCINE CONCERNS** |  |  |  |  |  |
| Negative experience/adverse reaction to vaccination in the past | 2 | 6 | 7 | 15 | 1.00 |
| Conspiracy theory | 0 | 0 | 5 | 5 | 0.33 |
| Fear of needles | 2 | 1 | 2 | 5 | 0.33 |
| Anti-vaccine in general | 0 | 0 | 4 | 4 | 0.27 |
| Opposing testing vaccines in animals | 0 | 1 | 0 | 1 | 0.07 |
|  | **4** | **8** | **18** | **30** | **2.00** |

